# Lower blood lactate and higher circulating natural killer cells at admission predict spontaneous survival in non-acetaminophen induced acute Liver failure

**DOI:** 10.1101/2020.11.27.20239731

**Authors:** Tanvi Agrawal, Rakhi Maiwall, Rajan V, Meenu Bajpai, Rakesh Kumar Jagdish, Shiv Kumar Sarin, Nirupma Trehanpati

**Affiliations:** Laboratory of Molecular Immunology, Department of Molecular and Cellular Medicine, Institute of Liver and Biliary Sciences New Delhi, India; Department of Hepatology, Institute of Liver and Biliary Sciences, New Delhi, India; Department of Transfusion Medicine, Institute of Liver and Biliary Sciences, New Delhi, India

**Keywords:** ALF, DAMPs, Immune cells, Lactate, NK cells

## Abstract

**Background and Aims:** Massive cellular necrosis in ALF is dominantly immune mediated and innate immune cells are major pathophysiological determinants in liver damage. Our aim was to investigate specific innate immune cells or damage associated molecular patterns (DAMPs) relating to the final outcome of patient.

**Methods:** In fifty ALF patients and in fifteen age-matched healthy controls (HC), DAMPs were measured in plasma using ELISA. Phenotypic analysis of neutrophils, monocytes, natural killer (NK) and NKT cells was done by flow-cytometry and correlated with clinical and biochemical parameters.

**Results:** ALF patients (aged 27±9 yr, 56% males, 78% viral etiology) had MELD of 31.5±8, jaundice to hepatic encephalopathy (HE) of 4.6±3.2 days, HE grade III-IV, 82% with cerebral edema, 38% met KCH criteria, 56% had suspected sepsis. Percentage of intermediate monocytes (CD14^+^CD16^+^) was increased (p<0.01) and non-classical monocytes (CD14^-^CD16^+^) was decreased in ALF compared to HC. CD16^+^CD56^+^ NK cells in total lymphocytes was significantly lower in ALF patients compared to HC, but was higher in survivors {9.28% (0.5-20.3)} than non-survivors {5.1% (0.2-10.6)} (p<0.001). Higher percentage of circulating NK cells (>6.7%) at admission was a good predictor of survival. Non-survivors had higher levels of serum lactate (6.1 vs. 28, Odds ratio 2.23, CI 1.27-3.94) and granzymeB positive NK cells than survivors. Logistic regression model predicted the combination of lactate levels with NK cell percentage at admission for survival (AUROC of 0.94; sensitivity 95.8%, specificity of 78.5%).

**Conclusion:** Combination of NK cell frequency and lactate levels at admission can reliably predict survival of ALF patients.

**KEY POINTS:**

- ALF is generally immune mediated and predominantly caused by viral infections or acetaminophen toxicity.
- Therapeutic options are limited in ALF, important to know key immune players for their survival.
- CD16^+^CD56^+^ NK cells were found to be higher in survivors than non survivors.
- Combination of lactate levels with NK cell percentage at the time of admission can reliably predict the survival of ALF patients.

## Introduction

Acute liver failure (ALF) is characterized by sudden loss of hepatic function due to excessive hepatocyte cell death in previously normal liver along with development of hepatic encephalopathy, coagulopathy, and jaundice, and can rapidly progress to multi-organ failure with high mortality rates (40-80%) [1, 2]. The predominant causes of ALF are viral infections in developing countries including India and drug-induced liver injury primarily acetaminophen toxicity in developed countries [3].

Although the pathophysiology of ALF is not fully understood, massive hepatocyte death beyond the liver regenerative capacity is considered as a core event in development of ALF and both direct damage and immune-mediated damage are implicated in this. ALF usually results in a systemic inflammatory response and progression to multi-organ failure. The systemic release of inflammatory mediators might be responsible for the progression from inflammatory response to multi-organ failure [4]. Necrotic or infected hepatocytes attract immune cells which release pro-inflammatory cytokines with subsequent hepatocellular injury [5]. Circulating neutrophils in ALF have been shown to have impaired bactericidal function, and this is likely to be relevant in the increased susceptibility to infection [6]. The resident hepatic macrophages (Kupffer cells) potentiate liver injury by sensing danger molecules and releasing pro- and anti-inflammatory mediators [7]. There is also evidence for tolerance of circulating monocytes to bacterial endotoxin, which further impedes host immunity [8]. Infiltrating monocytes can mature into monocyte-derived macrophages (MoMF), which in cooperation with neutrophils, also involved in the resolution of inflammation.

Dead, necrotic and damaged cells release damage associated molecular patterns (DAMPs) [9], which are recognized by resident hepatic macrophages, Kupffer cell (KC), and neutrophils, leading to the activation of these cells [10] and resultantly initiate and extend sterile inflammation significantly contributing to hepatotoxicity [9]. Elevated circulating levels of DAMPs in patients with severe acute pancreatitis (SAP) have shown association with disease severity [11]. Thus, the dysregulation of pro-and anti-inflammatory factors leads to uncontrolled inflammation and progressive liver damage beyond the regeneration capacity of the liver [12].

Therapeutic options are limited in ALF and often etiology related [13]. If specific therapy fails the only curative option is emergency liver transplantation, which is often not feasible. Further, in ALF cases it has been observed that survival does not depend on aetiology or severity and can have different outcomes in patients with same aetiology and severity. However, the factors responsible for helping a native liver to regenerate spontaneously are largely unknown.

Previous studies on factors responsible for prediction of spontaneous survival were largely dependent on biochemical parameters [14-20]. However, keeping in mind the key role played by innate immune cells in pathophysiology of ALF we thought it would be worthwhile to study the dynamics of innate immune cell population in circulation in ALF patients.

Thus, we designed this study with the aim to identify if the numbers of any particular innate cell population upon admission can determine the spontaneous survival of patient despite having any aetiology or degree of severity. We also studied damage associated molecular patterns (DAMPs) levels as they stimulate other innate cells and thus play an active role in outcome. The main aim of study was to identify innate immune cells which may play a larger role in survival ALF patients. It would also help in deciding the treatment strategy and intensity, i.e., maximum supportive therapy while waiting for the liver to regenerate or deciding to list the patient for liver transplantation [2].

## Material and Methods

### Patient enrolment

Acute liver failure patients admitted to Liver Coma Intensive Care Unit at the Institute of Liver and Biliary Sciences, New Delhi, India were enrolled from March 2017 to March 2019. Acute liver failure patients were defined as those with development of jaundice, coagulopathy (INR >1.5), and hepatic encephalopathy within 4 weeks of the onset of symptoms in a patient with a previously healthy liver. The eligibility criteria were age greater than 18 years and lower than 65 years. All non-acetaminophen aetiologies were included and patients with no consent given, any form of known chronic liver disease, having human Immunodeficiency virus (HIV) infection or patients who underwent transplant for liver failure were excluded. All patients underwent clinical examination and diagnostic screen. The protocol was approved by the institutional review board and ethics committee. Fifteen age and sex matched healthy controls with no prior history of any liver related disease and negative for Immunoglobulin M (IgM) hepatitis A virus (anti-HAV), hepatitis E virus (anti-HEV), hepatitis B surface antigen, anti-HCV and HIV were also enrolled in the study. Informed consent was obtained from each patient or their immediate family members and healthy controls. Each patient was followed either till discharge from hospital or death. Clinical parameters like age, gender, presentation (hyperacute/acute), Jaundice to hepatic encephalopathy (HE) duration, KCH criteria, HE grade, presence or absence of systemic inflammatory response syndrome (SIRS) were recorded.

### Sample collection

Peripheral blood samples (10-15 ml) were collected from all the patients at baseline (at the time of admission). Blood samples were analysed for laboratory parameters (AST, ALT, complete blood count (CBC), prothrombin time (PT), international normalized ratio (INR), bilirubin, creatinine, blood urea, lactate and arterial ammonia at the time of admission.

### Characterization of Neutrophils, Monocytes and Natural Killer cells

Fresh whole blood samples were stained for innate cell specific antibodies and were analysed for neutrophil, monocyte and NK markers. Whole blood samples were stained with anti-human CD14-FITC (fluorescein isothiocyanate), anti-human CD16-BV410 (Brilliant Violet 410) and anti-human HLADR-PECy7 (Phycoerythrin/cyanine 7) for monocytes. Neutrophils were identified using anti-human CD11b-PECy7 and anti-human CD16-BV410 along with anti-human CXCR1-APC (allophyacocyanin), anti-human CXCR2-FITC and anti-human CD66B-PE (Phycoerythrin). Natural Killer cells were identified using a cocktail of anti-human CD3-FITC and anti-human CD16/56-PE (Biolegend Corporation, USA).

In brief, 60µl whole blood was incubated at room temperature for 25 minutes with respective cocktails of antibodies for different cell types. Erythrocytes were lysed with 1x RBC lysis buffer. Remaining leucocytes were washed twice with phosphate buffer saline. Granzyme B expression in natural killer cells was studied by fixing and permeabilising the cells using cytofix/cytoperm (BD Biosciences, USA) and staining with anti-human granzyme B-APC antibody (BD Biosciences). A minimum of 1,00,000 events were acquired on FACS Verse (BD Biosciences) and data was analysed using FlowJo software (Flow Jo LLC, USA). Cells were identified and gated using forward and side-scatter characteristics. Percentage of immune cell population along with mean fluorescence intensity for various markers present in the blood sample was calculated.

### Measurement of DAMPs

Quantitative measurement of DAMPs [HMGB1 (High Mobility Group Box 1), heat shock protein 60 and 70 (HSP60, HSP70), Serum Amyloid A (SAA), S100A8, S100A9, Interleukin-33 (IL-33)] was done in plasma of ALF patients by commercially available Enzyme linked Immunosorbent Assay (ELISA) kits according to the manufacturer’s instructions (R&D Systems, MN, Canada for IL-33 and Kinesis Dx, CA, USA for other DAMPs). The minimal detectable limit of assays were (2ng/ml for HMGB1, 0.1ng/ml for HSP60, 0.5ng/ml for HSP70, 1µg/ml for SAA, 2.15ng/ml for S100A8, 1.47ng/ml for S100A9 and 1.65pg/ml for IL-33). All the assays were performed in duplicate.

### Statistical Analysis

The Mann–Whitney U test was used for comparing two groups. The results were presented with 95% confidence interval (CI) and P value >0.05 was considered significant. A receiver operating characteristic (ROC) curve was made using the outcome variable as death or survival and admission levels for various parameters as the independent variable for predicting mortality or survival. Sensitivity, specificity, and odds ratio (OR) for the cut off value were also derived. Univariate and multivariate analysis using linear regression was used to make model for prediction. Statistical software SPSS (version 10; SPSS Inc., Chicago, Illinois, USA) and STATA (version 8.0) was used for statistical analysis.

## Results

### Demographic profile

The demographic profile of patients is as shown in Table 1. A total of 52 ALF patients were enrolled in the study. Two patients underwent transplant and were thus excluded from the study leaving the total number of patients to 50. Seventy-four percent of cases had a viral aetiology. Hepatitis E infection was the most common aetiology (n=19, 37.2 %) followed by hepatitis A (n=13, 25.49%), hepatitis B (n=4, 7.8%), other viruses (n=3, 5.8%), non-acetaminophen drugs (n=3, 5.8%) and other causes (n=3, 5.8%). Aetiology could not be determined for 5 patients (9.8%). Thirty (60.0%) patients did not survive. HE grade of I-II was observed in 9 (17.64%) patients while rest of the patients had HE grade III-IV. Eight two percent patients had cerebral edema based on CT and 19 (38.0%) patients met King’s college hospital (KCH) criteria for poor prognosis. Infection was diagnosed in 28(56.0%) patients at the time of admission as proven by microbiological analysis of mini-brachoalveolar lavage.

**Table 1:** Demographic profile of patients at admission

| Parameter | HC | ALF (survivors) | ALF (non-survivors) | p value | OR (95% CI) |
| --- | --- | --- | --- | --- | --- |
| Patients, n (% of total) | 15 | 20 (40.00) | 30 (60) |  |  |
| Age in years | 33 (19-52) | 27 (18-48) | 23 (18-49) | 0.42 | 1.04 (0.95-1.13) |
| Sex M (%) | 6 (40.0) | 9 (40.0) | 19 (63.3) | 0.2 | 0.47 (0.15-1.50) |
| Etiology | NA |  |  |  |  |
| HEV |  | 7 | 12 |  |  |
| HAV |  | 10 | 3 |  |  |
| HBV |  | 1 | 3 |  |  |
| other Virus |  | 0 | 3 |  |  |
| drugs |  | 0 | 3 |  |  |
| Others |  | 2 | 3 |  |  |
| Indeterminate |  | 0 | 5 |  |  |
| MELD score | NA | 24 (17-49) | 34 (20-49) | **0.014** | 1.11 (1.02-1.21) |
| HE Grade | NA |  |  |  |  |
| I-II, n(%) |  | 6 (30.0) | 3 (10.0) |  |  |
| III-IV, n(%) |  | 14 (70.0) | 26 (86.6) | 0.15 | 2.79 (0.67-11.55) |
| Jaundice to HE Duration | NA | 4 (1-7) | 4 (1-18) | 0.31 | 1.11 (0.91-1.33) |
| Meeting KCH criteria, n(%) | NA | 4 (20.0) | 15 (50.0) | **0.032** | 4 (1.08-14.81) |
| Hyperacute liver failure | NA | 16 (80.0) | 18 (60.0) | 0.18 | 0.37 (0.1-1.39) |
| SIRS, n(%) | NA | 6 (30.0) | 14 (46.7) | 0.24 | 2.04 (0.62-6.75) |
| AKI, n(%) | NA | 7 (35.0) | 21 (70.0) | **0.015** | 4.33 (1.30-14.45) |
| Bilirubin (mg/dl) | NA | 12.4 (3.8-62) | 15.3 (3.8-49) | 0.83 | 1.01 (0.96-1.06) |
| INR | NA | 2.39 (1.3-3.6) | 3.98 (1.3-10.4) | **0.01** | 1.78 (1.15-2.75) |
| Creatinine (mg/dl) | NA | 0.56 (0.1-4.8) | 0.7 (0.02-4.96) | 0.68 | 1.11 (0.69-1.77) |
| Urea (mg/dl) | NA | 23 (8-209) | 16 (4.28-154) | 0.15 | 0.99 (0.68-1.01) |
| AST (IU/L) | NA | 896 (87-3563) | 268 (41-4211) | 0.71 | 1.00 (1.00-1.00) |
| ALT (IU/L) | NA | 501 (59-4447) | 1644 (24-5357) | 0.11 | 1.00 (0.99-1.00) |
| Ammonia (µg/dL) | NA | 218 (88-424) | 356 (117-761) | **0.002** | 1.01 (1.00-1.02) |
| Lactate (mmol/L) | NA | 2.8 (1.3-4.8) | 6.1 (1.3-18) | **0.005** | 2.23 (1.27-3.94) |
| Suspected Infection n(%) | NA | 11 (55.0) | 17 (56.7) | 0.91 | 1.07 (0.34-3.34) |

Data is presented as median. Figures in brackets indicate range or percentage. P value significant if less than 0.05
HEV-Hepatitis E virus; HAV-Hepatitis A virus; HBV-Hepatitis B virus; MELD-Model of End Stage Liver Disease; HE-Hepatic encephalopathy; KCH-King's College Hospital; SIRS-Systemic Inflammatory Response Syndrome; AKI-Acute Kidney Injury; INR-Internalized Normalized Ratio; AST-Aspartate aminotransferase; ALT-Alanine transaminase.

### Damage Associated Molecular Patterns

Levels of various DAMPs were measured at the time of admission in ALF patients. Median levels of DAMPs in plasma of ALF patients, (survivors and non-survivors) is as shown in Table 2. No significant difference in levels of DAMPs was noted between survivors and non-survivors. To see if the level of any DAMP at the time of admission predicted survival, a ROC curve of DAMPs of each patient as the independent variables and mortality as the outcome variable was made. DAMPs were found to be very poor predictors of mortality with area under the curves as shown in Figure 1.

**Table 2:**
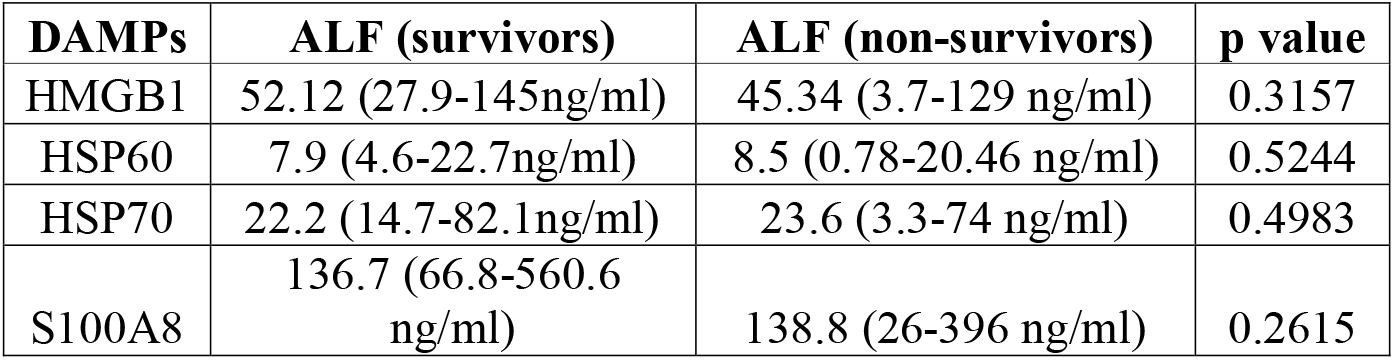
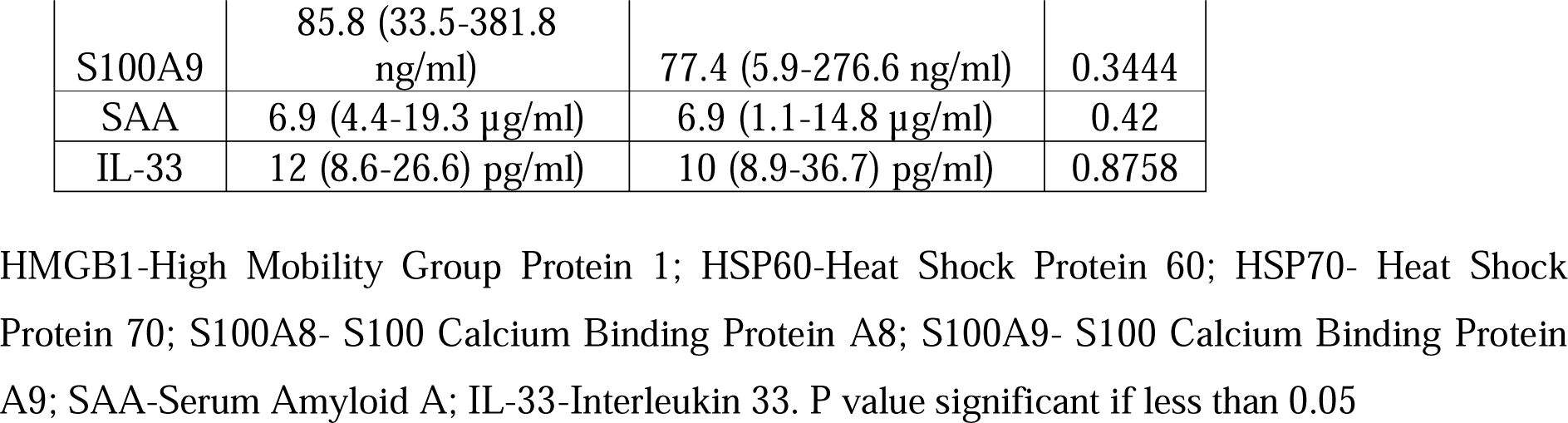
Median levels of DAMPs in plasma of ALF patients at the time of admission

| DAMPs | ALF (survivors) | ALF (non-survivors) | p value |
| --- | --- | --- | --- |
| HMGB1 | 52.12 (27.9-145ng/ml) | 45.34 (3.7-129 ng/ml) | 0.3157 |
| HSP60 | 7.9 (4.6-22.7ng/ml) | 8.5 (0.78-20.46 ng/ml) | 0.5244 |
| HSP70 | 22.2 (14.7-82.1ng/ml) | 23.6 (3.3-74 ng/ml) | 0.4983 |
| S100A8 | 136.7 (66.8-560.6 ng/ml) | 138.8 (26-396 ng/ml) | 0.2615 |
| S100A9 | 85.8 (33.5-381.8 ng/ml) | 77.4 (5.9-276.6 ng/ml) | 0.3444 |
| SAA | 6.9 (4.4-19.3 µg/ml) | 6.9 (1.1-14.8 µg/ml) | 0.42 |
| IL-33 | 12 (8.6-26.6 pg/ml) | 10 (8.9-36.7 pg/ml) | 0.8758 |

HMGB1-High Mobility Group Protein 1; HSP60-Heat Shock Protein 60; HSP70- Heat Shock Protein 70; S100A8- S100 Calcium Binding Protein A8; S100A9- S100 Calcium Binding Protein A9; SAA-Serum Amyloid A; IL-33-Interleukin 33. P value significant if less than 0.05

**Figure 1:**
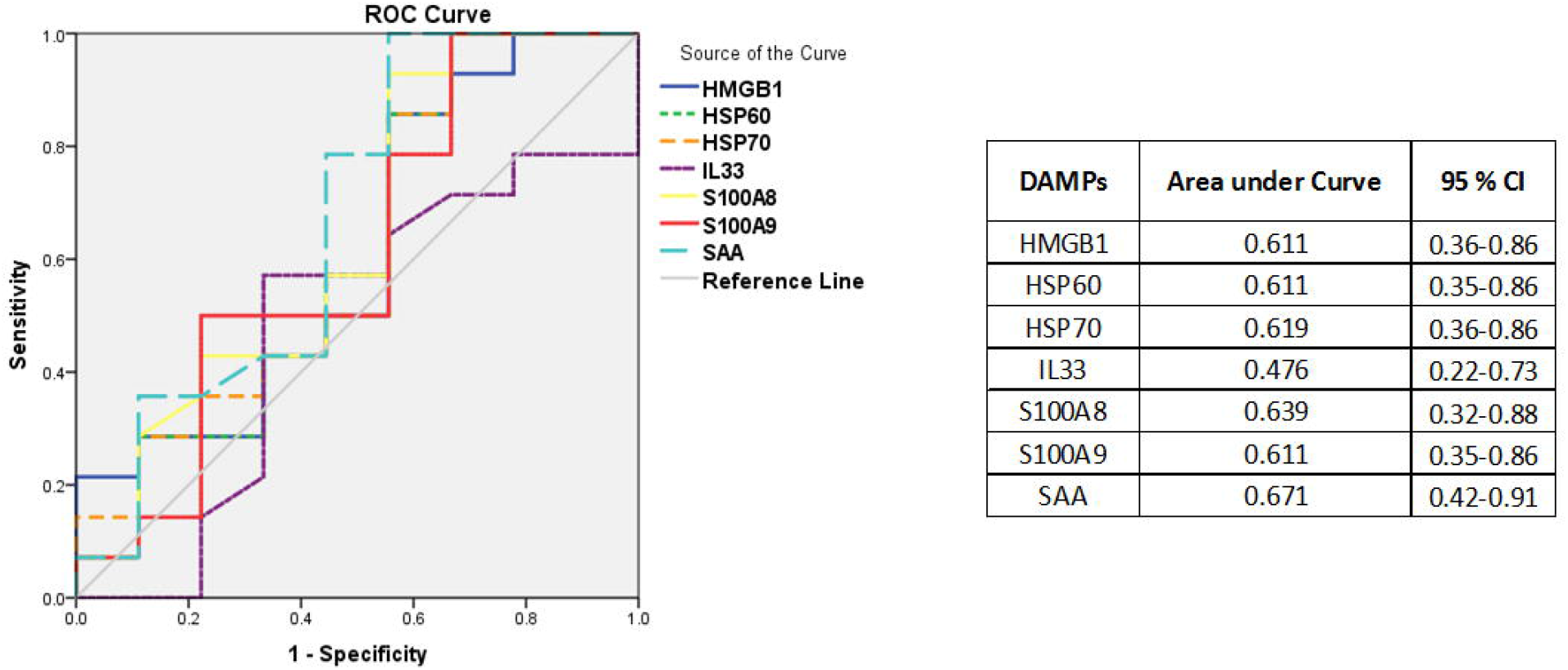
Receiver operating characteristic (ROC) curve of Damage Associated Molecular Patterns and mortality.

### Neutrophil, Monocyte and Natural Killer cell population in ALF patients

Characterization of neutrophils, monocytes and natural killer cells in whole blood is as shown in Supplementary figure 1. Neutrophils were identified as CD11B+CD16+ population among granulocytes. Based on the expression of CD14 and CD16 monocytes were characterised as either classical monocytes (CD14+CD16-/lo), intermediate monocytes (CD14+CD16+) and non-classical monocytes (CD14-/loCD16+). Natural Killer cells were identified as CD16+CD56+ population among lymphocytes while Natural Killer T cells were identified as CD16+CD56+CD3+ population. The percentage of CD16+CD56+ population among lymphocytes was considered as the percentage of circulating NK cells in all the results to maintain homogeneity as presence of red blood cells in processed sample can alter the actual percentage of NK cell present if considered in total population..

At the time of admission, no difference was observed in the number of neutrophils among HC and ALF patients, however, expression of CD66B on neutrophils was significantly higher (p<0.0001) while the expression of CXCR1 was significantly lower in ALF patients (p<0.0001) as compared to HC (Figure 2). As for monocyte population, no difference was observed in classical monocyte population however, intermediate monocytes were significantly higher in ALF patients while non-classical monocytes were significantly (p<0.0001) lower in ALF patients compared to controls at the time of admission. Natural killer and natural killer T cells were lower in ALF patients as compared to controls, however it was significant only for Natural Killer cells (p<0.01).

**Figure 2:**
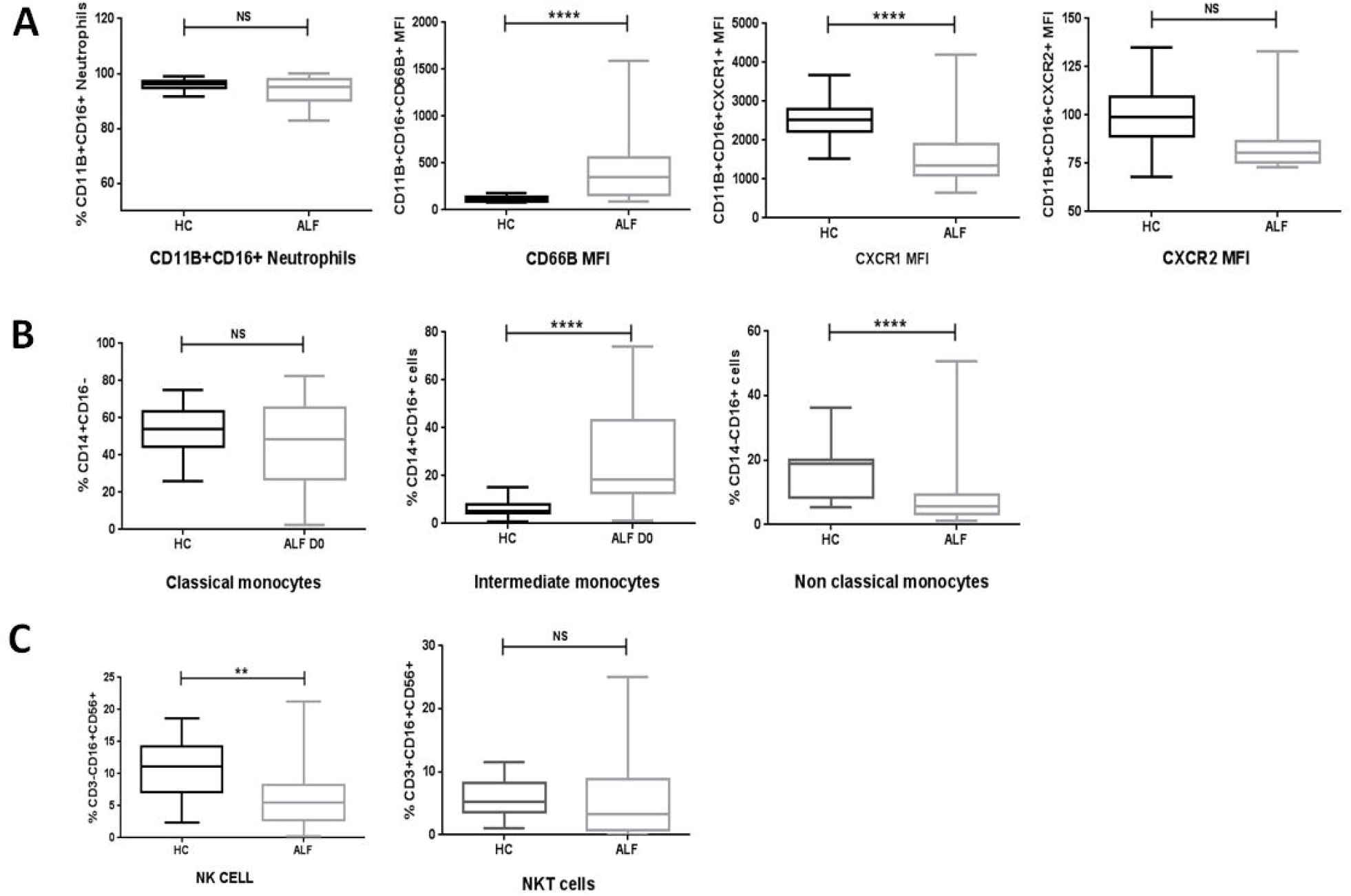
Percentage frequencies of Circulating (A) Neutrophils, (B) monocytes and (C) Natural Killer Cells in HC and ALF patients. Box plot representing with mean and SD. P value > 0.01 is represented as ** and > 0.001 is represented as ****

### Natural Killer cells were higher in ALF survivors versus non-survivors at admission and were predictive of survival

We further analysed the cell populations to see if there was any difference among survivors and non-survivors in any population. Neutrophils, monocytes and NKT cells did not show any difference between survivors and non-survivors (Supplementary figure 2). Circulating NK cells on the other hand were significantly higher in survivors as compared to non-survivors (p<0.001) (Figure 3A). The percentage of NK cells in survivors were comparable to those found in healthy controls. To study the cytotoxic capacity of NK cells we analysed and compared the percentage of granzyme B positive NK cells between survivors and non-survivors. As compared to survivors, NK cells from non survivors express significantly more granzyme B (p>0.05) (Figure 3B) suggesting that they are more cytotoxic.

**Figure 3:**
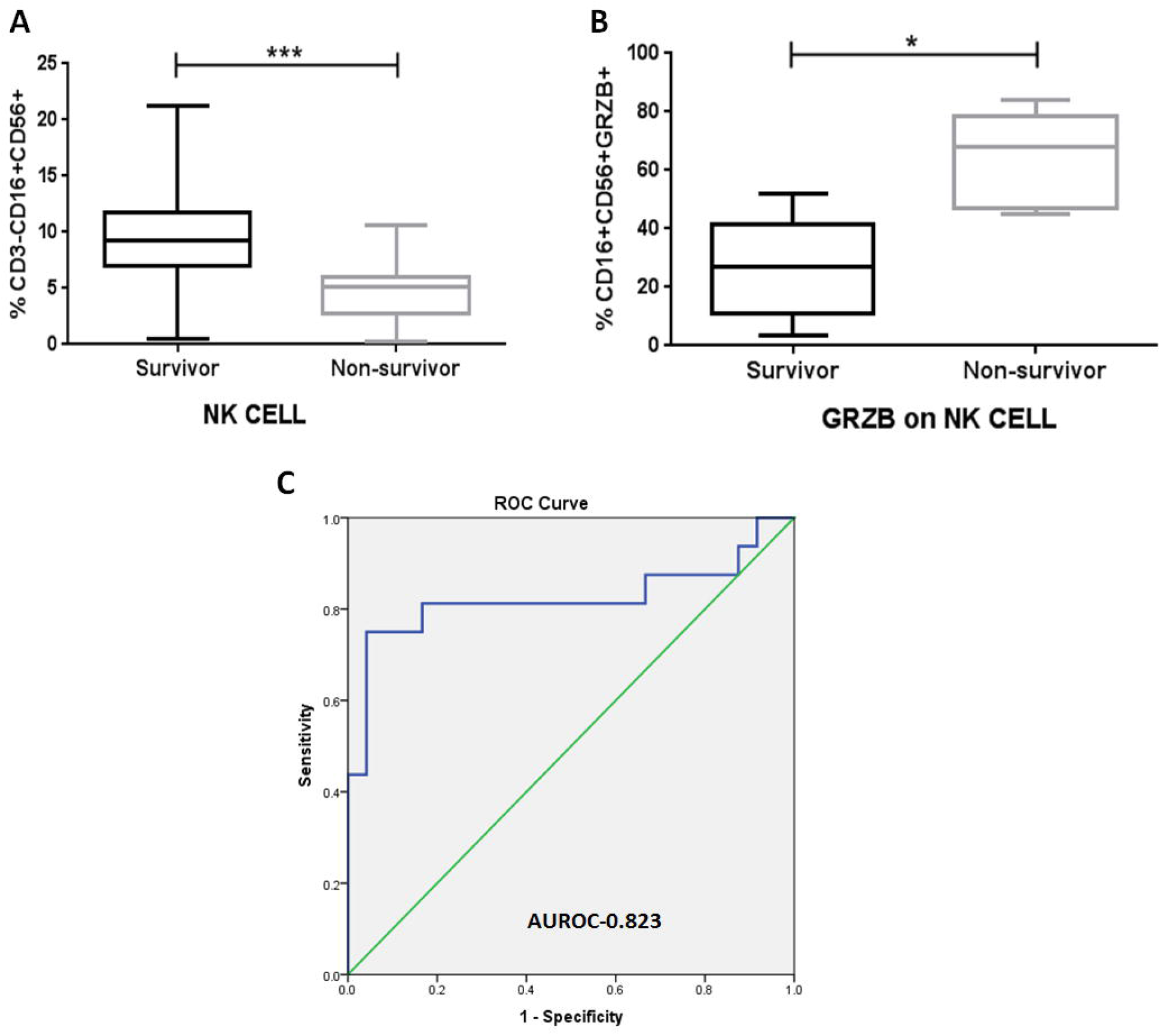
(A) Circulating peripheral NK cell percentage in ALF survivor’s vs non-survivors. (B) Expression of granzyme B on NK cells (C) Receiver Operating Curve (ROC) with NK cell percentage as the independent variable and survival as the outcome variable.

To check the prognostic predictive value of NK cell percentage a Receiver Operating Curve (ROC) was made for NK cell percentage of each patient at admission as the independent variable and survival as the outcome variable. The area under the ROC curve was 0.823 (95% CI 0.66–0.98) for NK cell percentage. An NK cell percentage of > 6.7 was found to be 81.3% sensitive and 83.3% specific for predicting survival (Figure 3C).

### Higher arterial ammonia, INR and arterial lactate were good predictors of mortality

Since previous studies have focussed on biochemical parameters for predicting survival we analysed these parameters in our patients. When compared between survivors and non-survivors upon univariate analysis it was observed that at admission, ammonia levels, INR, MELD score, AKI, and lactate levels were significantly different between survivors and non-survivors (Table 1) with median ammonia levels, INR, MELD score, and lactate levels being significantly higher in non-survivors as compared to survivors (p<0.01) (Figure 4). No differences in the levels of bilirubin, AST, ALT, urea or creatinine were observed when compared between survivors and non-survivors. A Receiver Operating Curve (ROC) with KCH for poor prognosis, ammonia levels, INR, MELD score and lactate levels of each patient as the independent variable and mortality as the outcome variable was made (Figure 5). The area under the ROC curve was 0.794 (95% CI 0.66– 0.92) for arterial ammonia, 0.77 (95% CI 0.62–0.91) for INR and 0.85 (95% CI 0.73–0.96) for lactate levels. An arterial ammonia level of >270 µg/dl was found to be 70.0% sensitive and 72.2% specific for predicting mortality while an INR of >2.6 was found to be 74% sensitive and 72.2% specific for predicting mortality. Lactate levels >3.3 was found to be 77.8% specific and 77.8% specific. KCH proved to be a very poor predictor as area under the curve was 0.59 (95% CI 0.42– 0.76).

**Figure 4:**
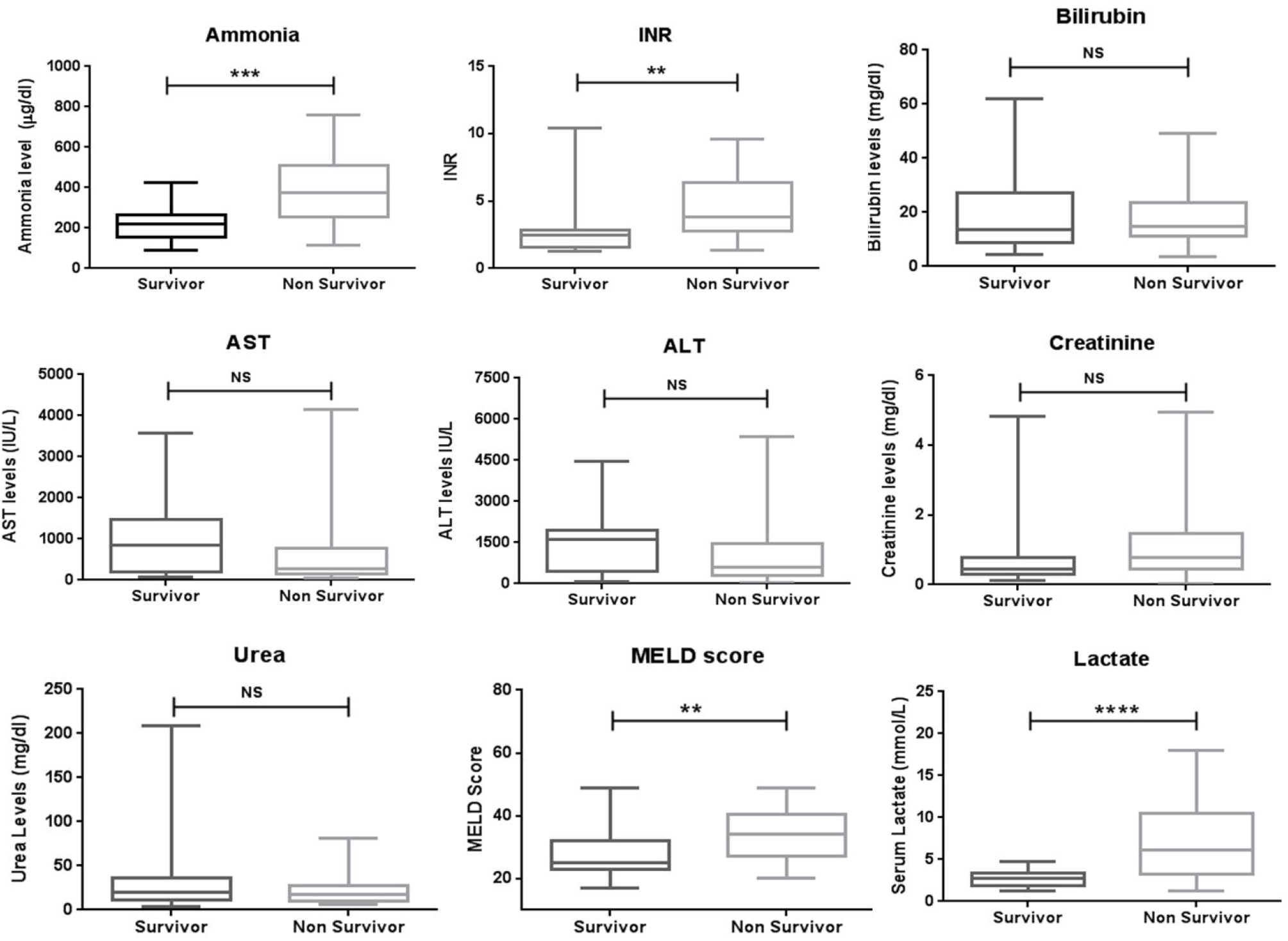
Levels of various biochemical parameters at the time of admission in ALF patients, who survived and not survived after a week.

**Figure 5:**
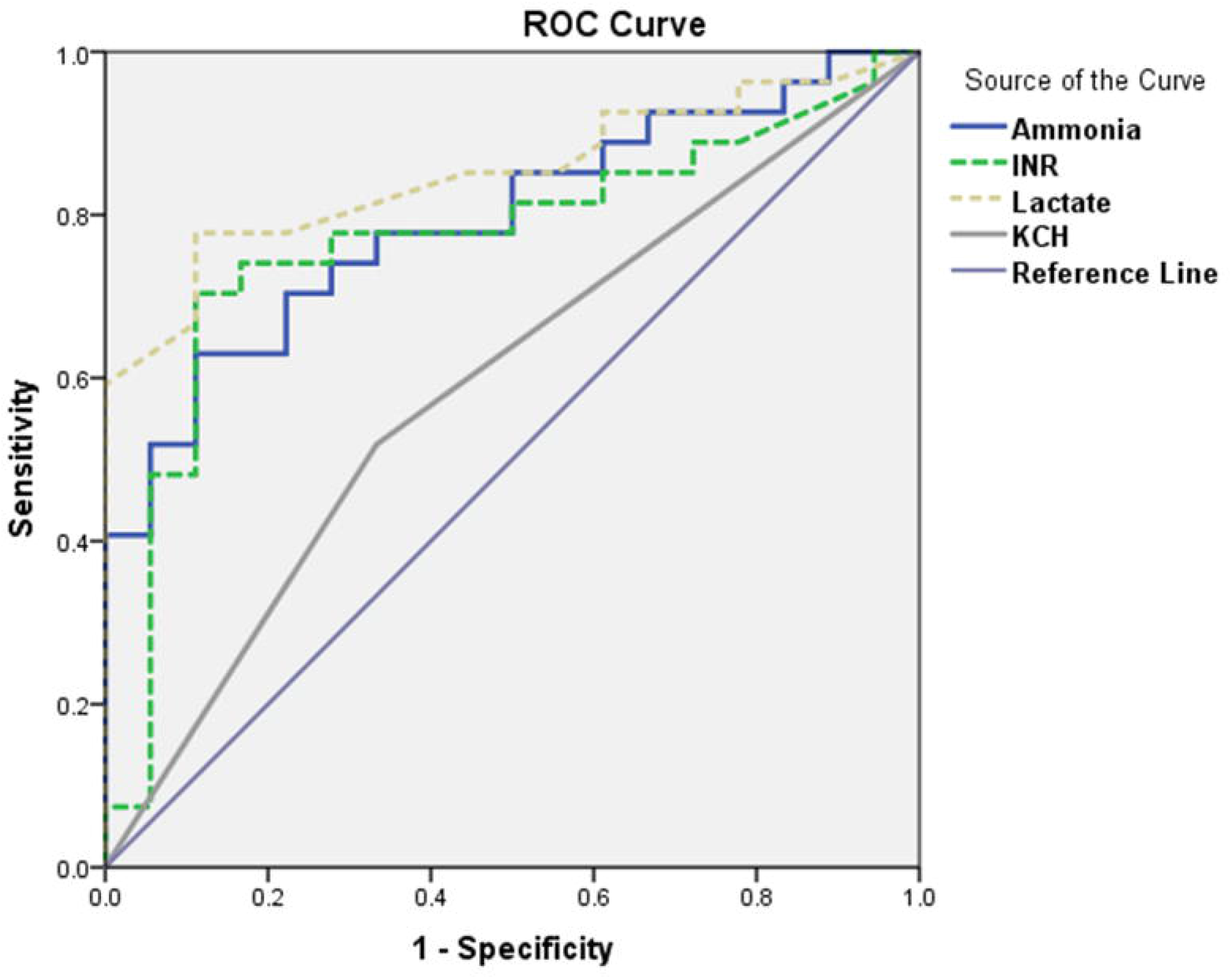
Receiver operating characteristic (ROC) curve of KCH criteria, arterial ammonia levels, INR and lactate at admission and mortality.

### Combination of blood lactate and NK cell percentage is best predictor of spontaneous survival

Since univariate predictive value of ammonia, INR, lactate and NK cell population were significant we tried to find whether a combination of these factors with NK cell percentage has increased predictive value for prognosis. Multiple logistic regression analysis of all these factors was done and it was observed that a combination of lactate and NK cell population was the best predictor of spontaneous survival. A logistic regression ROC curve was then made (Figure 6). The area under the curve was 0.943 (95% CI). The lactate-NK model has a sensitivity of 95.83%, specificity of 78.57%, positive predictive value of 88.46% and a negative predictive value of 91.67.

**Figure 6:**
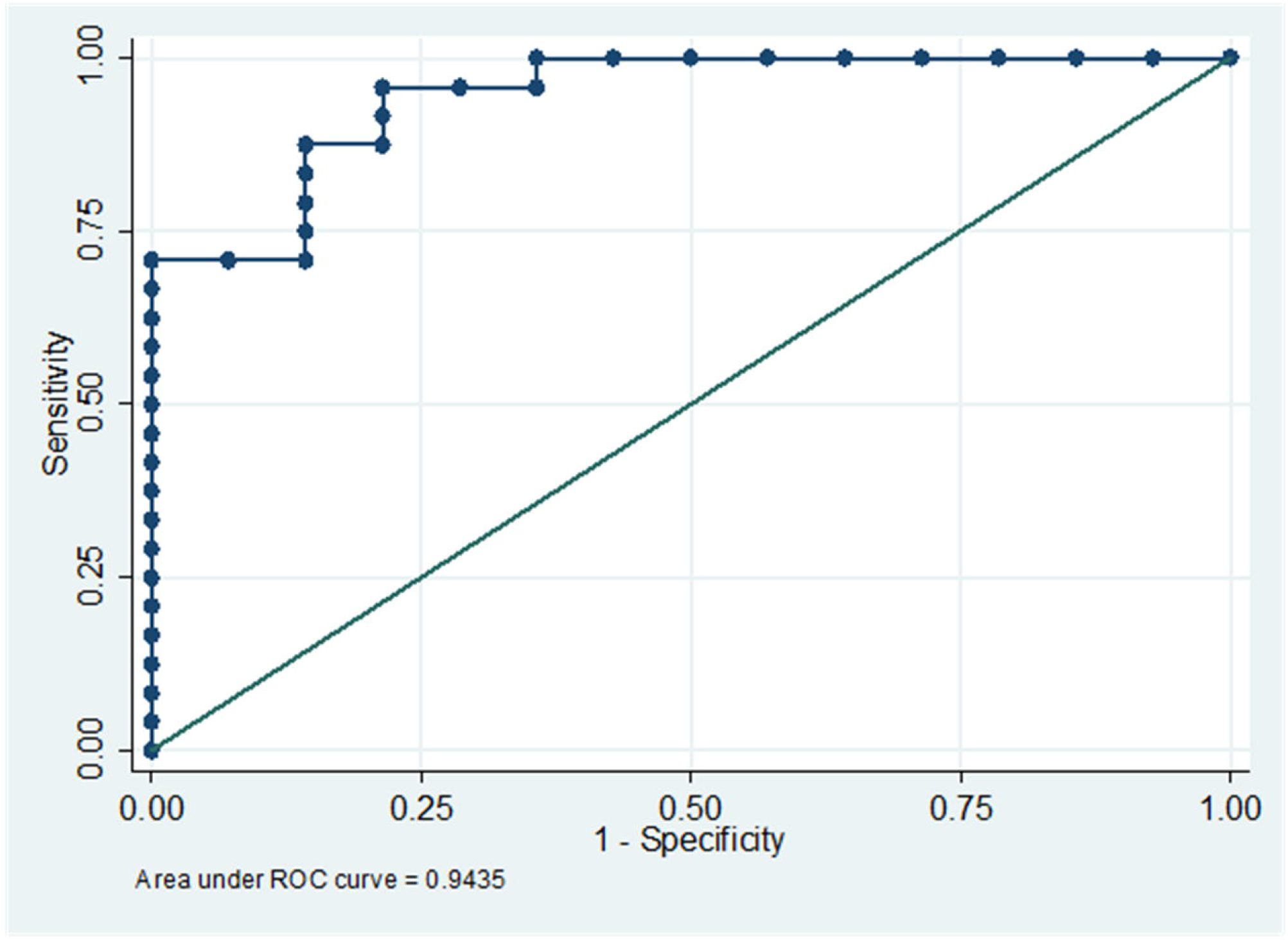
Logistic regression Receiver Operating Curve (ROC) with lactate and NK cell percentage taken as score as the independent variable and survival as the outcome variable.

## Discussion

We focussed our study on circulating innate immune cell population as pathophysiology of ALF largely depends on them. Our results show that the percentage of NK cells (CD16+/CD56+ cells in total lymphocytes) was significantly lower in ALF patients compared to healthy controls, and more so in non-survivors than survivors (p<0.001). The percentage of intermediate monocytes (CD14+CD16+) was increased (p<0.01) while that of non-classical monocytes (CD14-CD16+) was decreased in ALF compared to controls, though the difference was not significant between survivors. These data indicate that defective functioning of the cellular components in particular of the innate immune system are closely implicated in the outcome of ALF patients. This dysfunction of innate immune cells could possibly be related to increased risk of infection in patients with ALF and decreased survival.

Our study shows that percentage of circulating Natural Killer cells in ALF patients are significantly lower compared to healthy controls. Previous studies with either acetaminophen related ALF [21] or acute viral hepatitis E [22] have also shown reduced numbers of circulating NK cells in patients as compared to controls. However, no differences were seen in context of the outcome in ALF [21]. This reduction in the numbers of NK cells in circulation can be attributed to their movement to intrahepatic compartments as liver being the main disease site. In our study we also found that the percentage of NK cells in most of the survivors were equivalent to what were observed in healthy controls. Further, NK cell percentage in survivors was significantly higher at baseline compared to non-survivors and was the single best predictor of spontaneous survival among all parameters tested. This suggests that more NK cells are recruited to the liver in non-survivors and which might be more damaging to liver.

The liver is highly enriched in innate immune cells with NK cells being a main effector lymphocyte population [23]. Upon activation, NK cells produce cytokines and can induce apoptosis of target cells via release of perforin and granzyme [24]. The exact role of NK cells in liver diseases is not elucidated very well as there are contradicting studies. NK cell cytotoxicity has been shown to contribute to liver damage in different forms of liver diseases [25]. It has been shown that NK cells are activated and cytotoxic post partial hepatectomy and support hepatocellular proliferation boosting liver regeneration [26], however, some other studies suggest that NK cells negatively regulate liver regeneration and cause tissue damage [23]. In case of ALF, NK cells have been earlier shown to enhance acetaminophen (APAP)-induced liver damage in mice [27] and removal of NK cells improved outcome. To check the cytotoxic status of circulating NK cells we studied granzyme B positivity in NK population in blood. We observed that granzyme B positive NK cells were significantly higher in non-survivors although NK being less in numbers in these cases. Thus, it can be suggested that in patients with adverse outcome more NK cells are recruited to the liver and these NK cells are more cytotoxic leading to enhanced liver damage causing mortality. Since liver biopsies were not available confirmation of the above hypothesis remains to be warranted. The exact mechanism of why NK cells are more cytotoxic in non-survivors needs to be elucidated.

Since previous studies focussed on biochemical parameters for studying spontaneous survival we tried to find out whether combining NK cell number to any biochemical parameter can give us more clear detail of the outcome. Our results show that high ammonia and arterial blood lactate levels at admission were best predictors of mortality among biochemical parameters. A study by Bhatia et al., [28] had also shown that in patients presenting with viral aetiologies non-survivors had significantly higher median ammonia levels than survivors and the levels were predictive of outcome and can be used for risk stratification. In contrast to our results, a study by Taura et al [29] has shown that hyperlactatemia is not a prognostic factor in non-acetaminophen related acute liver failure. Keeping the above results in focus we tested whether a combination of ammonia, INR, lactate and circulating NK cell percentage has better prognosis. We found that using logistic regression the prognostic efficiency of combined baseline lactate and NK cell population was higher than any individual component.

In addition, our results also showed that in neutrophils degranulation marker CD66B was significantly increased in ALF patients, however, the migration marker CXCR1 was significantly reduced. Although reduction in CXCR1 has already been shown during acute inflammatory conditions this was the first time CD66B expression was studied in ALF. However, we could not find any difference in the expression of CD66B or CXCR1 between survivors and non-survivors. In monocytes, intermediate monocytes were significantly higher in ALF patients while non-classical monocytes were significantly lower though the difference was not significant between survivors and non-survivors. The same results have also been shown by Abeles et al., [30] who showed an increase in intermediate while a decrease in non-classical monocytes in ALF patients. Although, monocytes play a major role in inflammation their percentage does not show any correlation to survival. Since DAMP molecules released from damaged and necrotic hepatocytes may serve as a crucial link between the initial hepatocyte damage and the activation of innate immune cells following acute liver failure [31] we tried to assess if any DAMP molecule can predict massive liver damage and thus aid in predicting mortality. We found no significant differences in the level of any DAMPs between survivors and non-survivors and all of them had very little predictive value.

In conclusion, our results suggest that more recruitment of NK cells to the liver is somehow responsible for the failure of the native liver to regenerate leading to death. Although more specific studies are warranted to confirm this, but a correlation between lower number of NK cells in circulation in any aetiology or severity with death as outcome suggests a critical role of NK cells in ALF. The main limitation of our study is low number of patients and we are further validating our data with more patients to help us reach a final consensus. Finally, at the stage of admission only, NK cells along with blood lactate levels can help in identification of patients who could survive spontaneously. This would save on unnecessary liver transplants and would help in accurate identification of ALF patients who will not survive without a liver transplant.

## Data Availability

yes

## Author contribution

TA collected and processed all the samples, analysed the data and written the manuscript. RM provided clinical inputs, selected patients to be enrolled and reviewed the manuscript. RV, MB and RKJ assisted in patient and control enrolment. NT and SKS conceptualised the study, analysed data, provided inputs and critically reviewed the manuscript.

## Acknowledgement

The authors wish to acknowledge the staff of Molecular and Cellular Immunology Department who helped in sample collection.

