## Supplementary information for "Lower blood lactate and higher circulating natural killer cells at admission predict spontaneous survival in non-acetaminophen induced acute Liver failure"

**Supplementary Figure 1: Gating Strategy for identification of (A) neutrophils, (B) monocytes and (C) NK and NKT cells.**

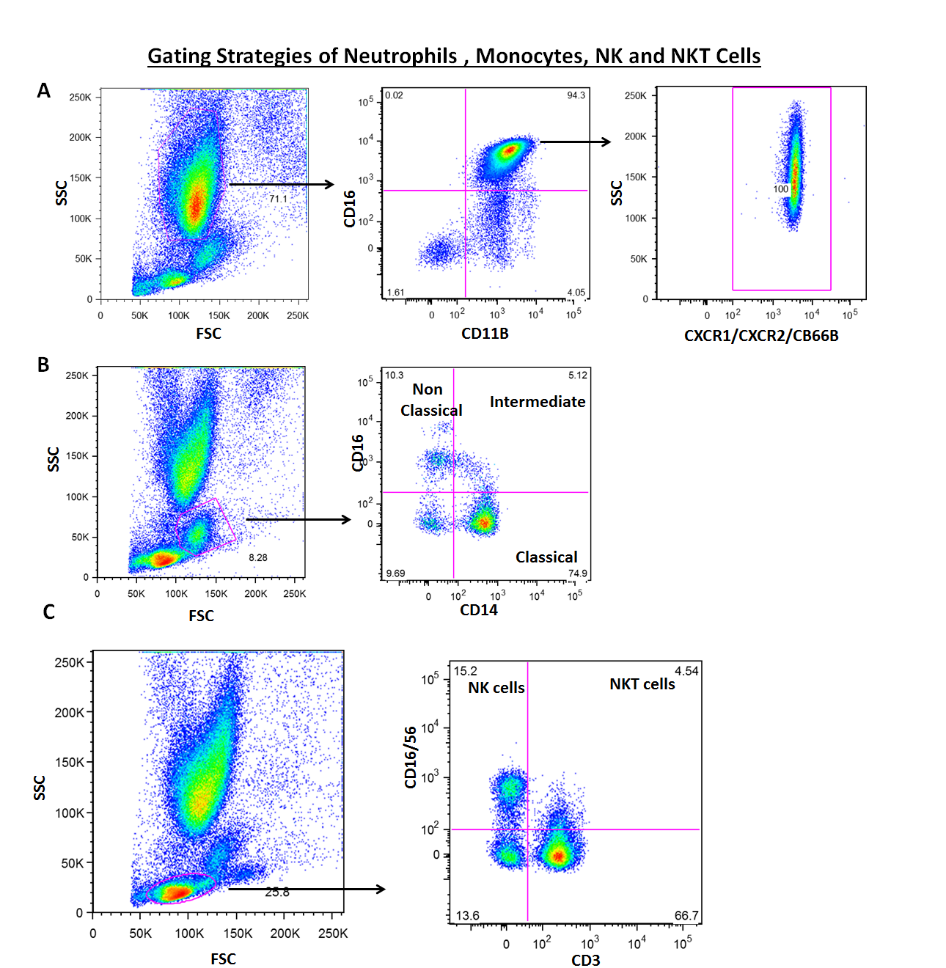

**Supplementary Figure 2: Population of circulating (A) Neutrophils, (B) monocytes and (C) Natural Killer T cells in survivors and non-survivors.**

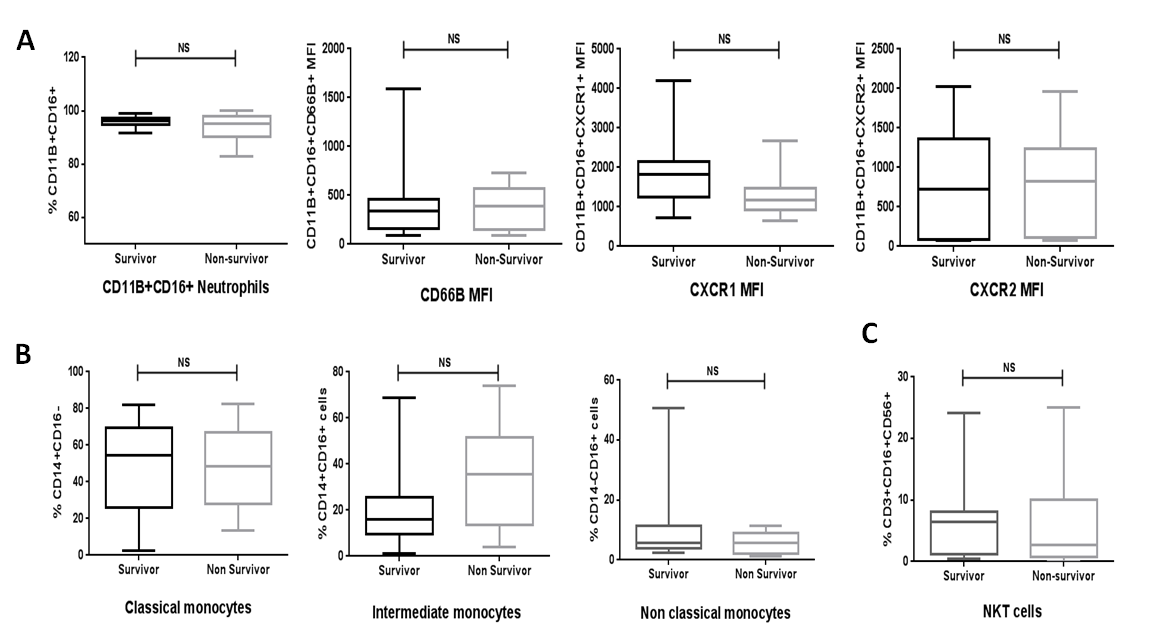
